# No evidence of increase in suicide in Greece during the first wave of Covid-19

**DOI:** 10.1101/2020.11.13.20231571

**Authors:** Sotiris Vandoros, Olga Theodorikakou, Kyriakos Katsadoros, Dimitra Zafeiropoulou, Ichiro Kawachi

**Affiliations:** King’s College London, London, UK; Harvard T.H. Chan School of Public Health, Boston, USA; Suicide Prevention Center, KLIMAKA NGO, Athens, Greece

**Keywords:** Covid-19, suicide, lockdown, unemployment, recession, excess mortality

## Abstract

**Background and Objective:** Mental health outcomes have reportedly worsened in several countries during the Covid-19 pandemic and associated lockdowns. In the present study we examined whether suicides increased in Greece during the first wave of the pandemic.

**Methods:** We used daily suicide estimates from a Suicide Observatory in Greece from 2015-2020 and followed three methodologies: A descriptive approach, an interrupted time series analysis, and a differences-in-differences econometric model.

**Results:** We did not find any empirical evidence of any increase in suicides during the first wave of Covid-19 and the lockdown in any of the three approaches used.

**Conclusions:** Suicides did not seem to increase during the first wave of covid-19 and lockdown in Greece. However, this does not mean that mental health did not deteriorate, or that we will not observe an increase in suicides during the second wave. Protective factors for Greece during the first wave may include working from home (for those able to tele-work), strong family ties, advertising of a suicide hotline and income support for the unemployed.

## 1. Background

In response to the first wave of the Covid-19 pandemic, countries throughout the world implemented lockdowns or other restrictions to slow down the spread of disease. These have had a devastating effect on economic activity, with rising unemployment rates and shrinking national income.^1-2^ Excess mortality during the Covid-19 pandemic has attracted much attention in the literature. Even disregarding deaths officially attributed to Covid-19, in many countries there are still excess deaths during the pandemic compared to what would have been expected considering mortality trends in previous years.^3-5^ While such evidence exists for a number of countries, this is not necessarily universal.

While data on the causes of other deaths are released with a significant time lag, there are possible spill-over effects of Covid-19 on other types of mortality, and one of the reasons that contribute to excess mortality may include suicides. However, there are counterarguments for decreased non-Covid-19 deaths, such as the possibility of fewer car crashes, work-related accidents and deaths from flu.

Financial hardship is associated with mental health problems,^6-8^ and events at the national level can also affect mental health and wellbeing.^9-10^ Increased suicide rates are often observed during recessions or times of rising unemployment,^11-15^ although some studies have found little or no evidence of a positive link between recessions and suicide.^16-18^ Economic uncertainty has risen to unprecedented levels during the Covid-19 pandemic,^19-20^ and previous evidence suggests that economic uncertainty is associated with increased suicide rates.^21^

Mental health has deteriorated during the pandemic due to the combined effects of financial insecurity and social isolation,^22-23^ and empirical evidence suggests that there is a bidirectional relationship between psychiatric conditions and Covid-19.^24^ Recent findings show that a quarter of American young adults contemplated suicide during this period,^25^ and there is evidence of an increase in suicide during the first wave of Covid-19 in Japan,^26^ which followed a decrease in suicides during the first wave.^27^ Previously, the SARS outbreak was associated with higher suicide rates in older adults.^28^

On one hand, there are factors that may lead to an increase in suicides during the pandemic. Unemployment increased sharply during the pandemic, and remote working can disrupt industries, with many people facing uncertainty about their future employment prospects.^11-15,21^ Even for those who were able to keep their jobs, furloughs and reduced working hours may have exacerbated financial insecurity. On the other hand, there are also factors that might put downward pressure on suicide rates. For those able to work remotely, the lockdowns may have meant avoiding a stressful work environment^29^ or commuting to work. Individuals may also save time from travelling to work, which would allow them to be involved in other activities such as exercising or spending time with family. By contrast, for those unable to tele-commute, the pandemic has been associated with constant worries about their own health or that of loved ones. Others have struggled to cope with the grief of losing loved ones to Covid-19, made even more difficult by social distancing policies. Lockdowns intensified social isolation and loneliness, resulting in a deterioration of mental health for the most vulnerable groups in the population.

The objective of this study is to examine whether there was an increase in suicides in Greece during the first wave of the Covid-19 pandemic and the associated lockdown. The study contributes to the literature by providing evidence on the association between the Covid-19 pandemic and suicide rates while the crisis unfolds, thus helping understand the spillover effects of the pandemic.

## 2. Data and Methods

The Centre for Suicide Prevention of NGO Klimaka has developed the Suicide Observatory, which records suicides in Greece on a daily basis. Klimaka is a recognised NGO that works closely with the Greek Ministry of Health and other public sector organisations. Suicide data are collected from a variety of sources, including coroners, funeral directors, prisons, relatives of people who have committed suicide, calls to the suicide prevention hotline, a network of volunteers (including health professionals, people registered with the Centre and civilians), and the electronic and print media. These data are not official, and they are generally incomplete. However, official data in Greece and elsewhere (including the US and the UK) are released with a significant time lag of about 2-3 years. As the Covid-19 pandemic unfolds and local or national lockdowns apply, it is important to provide swift evidence on the impact on suicide, to help design appropriate policies. Therefore, in the first instance we rely on the data provided by the Suicide Prevention Centre. In any case, we used the data from the Observatory for the entire study period, so we compared like with like, rather than using partial data from two different sources. Any bias in the post-Covid-19 period would also apply to the pre-covid-19 period, thus allowing us to study possible changes in suicide trends. Monthly unemployment rates were obtained from the Greek Statistics Authority.

The first Covid-19 case in Greece was reported on 26 February 2020, and the first death occurred on 12 March. The country went into a national lockdown on 23 March, which was lifted on 4 May. The number of cases and deaths remained relatively low during the summer of 2020.^30^

We distinguished between the pre-lockdown period and the post-lockdown period, thus using 23 March as the time of the intervention or the “treatment”. We then used two different “treatment” period lengths. In the first case, we used data up to 3 May, the last day of the lockdown, to restrict any effects to the duration of the actual lockdown, despite the ongoing pandemic. In the second case, we used data up to the end of August 2020, thus including observations after the lockdown, when, however, the Covid-19 problem remained, despite the lower infection and death rates. These two separate analyses would allow us to observe a broader picture on suicide trends during the pandemic in general. The very first death occurred just a few days before the national lockdown, and the authorities were swift in introducing measures, so the beginning of the Covid-19 problem in Greece roughly coincided with the lockdown.

We used three methodological approaches. First, we conducted a descriptive analysis, comparing the change in suicides in 2020 prior to and after the lockdown (i.e. before and after 23 March) to the corresponding figures in previous years.

Second, we conducted an interrupted time series (ITS) analysis to study any changes in suicide trends before and after the introduction of the first lockdown. The treatment period starts with the introduction of the lockdown, on 23 March 2020.

Third, we used a difference-in-differences (D-I-D) econometrics approach, which requires a control group. Covid-19 has affected nearly all aspects of everyday life worldwide, making it impossible to identify a population that has not been exposed to the treatment. Therefore, we used previous calendar years as a control group. We would expect that in the absence of the pandemic, *differences* in suicide rates between the period prior to 23 March and the period after 23 March would not differ compared to the corresponding differences over the same period in previous calendar years. This approach as previously been followed in previous studies.^3,9,31^ We used daily suicides from 2015 to 2019 as a control for suicides in 2020. The treatment group is year 2020, while control groups are the previous calendar years. The treatment period variable is the period 23 March onwards. The main variable of interest is the interaction between the two (treatment group * treatment period), which provides the difference-in-differences interaction term. This model is subject to an assumption about the common trend, which is hard to confirm given the daily nature of our data, which is why the D-I-D analysis is used as an additional check.

We estimated two sets of empirical models, with and without controlling for unemployment, as it has long been associated with suicides,^11-15^ and to filter out financial conditions from the effect of lockdowns and Covid-19. We also controlled for seasonality using month dummy variables; for day of the week effects using day of the week dummies; and for annual trends using year dummies.

## 3. Results

Figure 1 shows the average number of daily suicides in the periods from 1 January to 23 March 2020 (before the lockdown) and from 24 March to 3 May in 2020 (the lockdown period); compared to the previous 5 years (2015-2019). Panel A shows the average of 2015-2019 compared to 2020, while Panel B shows a detailed breakdown by year. As the graph shows, even before Covid-19 there was an upward trend in suicides annually, which might reflect the financial situation of Greece as it emerged from a long recession.^32^ In every single year between 2015 and 2019, in the period from 24 March to 3 May there was an absolute increase in the daily number of suicides compared to the period from 1 January to 23 March (on average from 0.97 to 1.17 – which constitutes a 20.13% increase). However, in 2020 there was a slight decrease (from 1.33 per day to 1.31 per day - a 1.49% decrease), and the “spring time excess” in suicides that was demonstrated in previous years seems to have been dampened during the first wave of the pandemic. Therefore, graphically, there is no evidence of an increase in suicides after the introduction of the lockdown, which also followed just a few days after the first Covid-19 death in the country.

**Figure 1.**
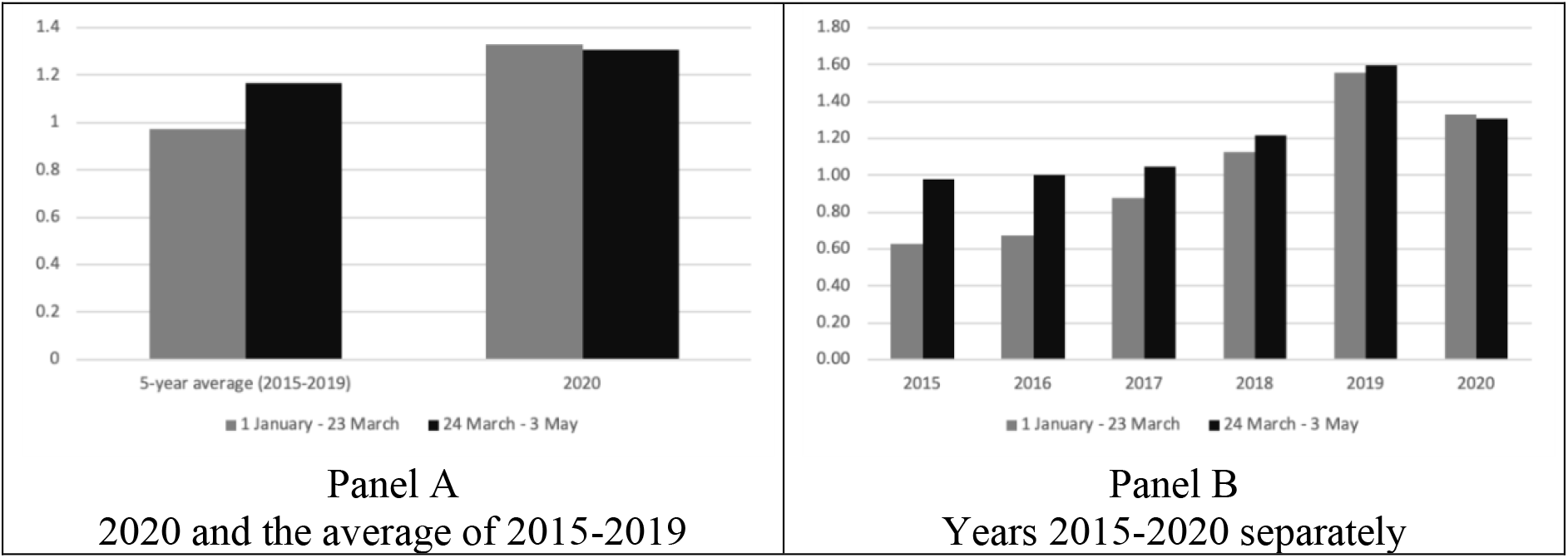
Average number of daily suicides before and after the introduction of the lockdown on 24 March 2020, up to the end of the lockdown on 3 May 2020. Data source: Suicide Observatory, Suicide Prevention Centre, Klimaka NGO

Figure A1 in the Appendix shows daily suicide rates from 1 January to 31 August, thus including the period after the lockdown was lifted. In this case, suicides in 2020 demonstrate an increase post-23 March compared to the period from 1 January to 22 March (from 1.33 to 1.45 per day – a 9.13% increase). However, the increase is smaller than that in previous years (from 0.97 to 1.28 per day – a 31.80% increase).

Results of the interrupted time series analysis are presented in Table 1. Column 1 shows the basic model without controlling for any other variables. Our findings show that there might be a decrease in the absolute level of suicides, although statistically significant only at the 10% level [coeff: -0.60; 95%CI: -1.309 to 0.111]. There is no statistically significant change in the trend of suicides [coeff: 0.02; 95%CI: -0.013 to 0.047]. Column 2 presents results when controlling for unemployment as well as day of the week, monthly and year effects. There is no statistically significant change in the absolute level [coeff: -0.52; 95%CI: -1.269 to 0.235] or trend [coeff: 0.02; 95%CI: -0.013 to 0.049] of suicides compared to the pre-lockdown period.

**Table 1.**
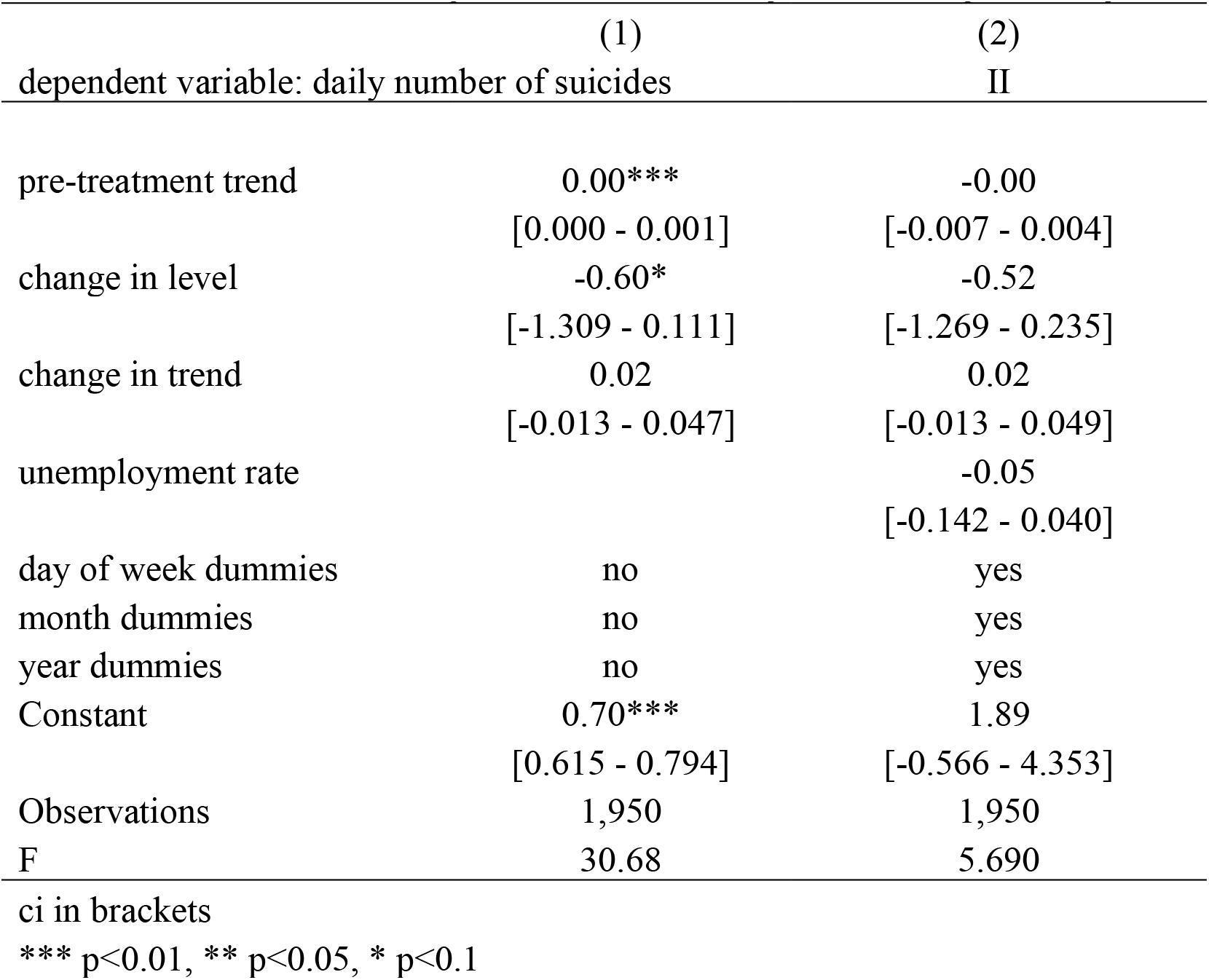
Results of the Interrupted Time Series analysis. Period up to 3 May 2020

|  | (1) | (2) |
| --- | --- | --- |
| dependent variable: daily number of suicides |  | II |
| pre-treatment trend | 0.00***<br>[0.000 - 0.001] | -0.00<br>[-0.007 - 0.004] |
| change in level | -0.60*<br>[-1.309 - 0.111] | -0.52<br>[-1.269 - 0.235] |
| change in trend | 0.02<br>[-0.013 - 0.047] | 0.02<br>[-0.013 - 0.049] |
| unemployment rate |  | -0.05<br>[-0.142 - 0.040] |
| day of week dummies | no | yes |
| month dummies | no | yes |
| year dummies | no | yes |
| Constant | 0.70***<br>[0.615 - 0.794] | 1.89<br>[-0.566 - 4.353] |
| Observations | 1,950 | 1,950 |
| F | 30.68 | 5.690 |

ci in brackets
\*\*\* p<0.01, \*\* p<0.05, \* p<0.1

Apart from studying the period up to the end of the lockdown (3 May), we also extended the study period to subsequent months, up to 31 August. Table A2 in the Appendix presents results of the interrupted time series for this extended period. Once again, we find no statistically significant change in the absolute level [coeff: -0.18; 95%CI: -0.619 to 0.255] or trend [coeff: 0.0005; 95%CI: -0.0039 to 0.0049] of suicides in the basic model. When including control variables we again did not find any increase in the absolute level [coeff: -0.154; 95%CI: -0.670 to 0.363] or trend [coeff: 0.001; 95%CI: -0.004 to 0.006].

Results of the differences-in-differences model are provided in Table 2. Column 1 presents the model without any control variables, while column 2 provides the full model, including the unemployment rate and day of the week, month and year dummies. In the basic model (column 1), the coefficient of the difference-in-differences interaction term is statistically insignificant, indicating that trends in suicides did not change after the introduction of the lockdown compared to trends in previous years [coeff: -0.22; 95%CI: -0.727 to 0.295]. Results persist when considering the full model in column 2 [coeff: -0.15; 95%CI: -0.675 to 0.365].

**Table 2.** Results of the difference-in-differences model. Period 1 January – 3 May of each calendar year

|  | (1) | (2) |
| --- | --- | --- |
| dependent variable: daily number of suicides |  |  |
| diff-in-diff interaction term | -0.22<br>[-0.727 - 0.295] | -0.15<br>[-0.675 - 0.365] |
| treatment period (24 March - 3 May) | 0.20**<br>[0.002 - 0.391] | 0.17<br>[-0.234 - 0.565] |
| treatment group (year 2020) | 0.36***<br>[0.095 - 0.623] | -0.42<br>[-1.680 - 0.849] |
| unemployment rate |  | -0.11*<br>[-0.227 - 0.016] |
| day of week dummies | no | yes |
| month dummies | no | yes |
| year dummies | no | yes |
| Constant | 0.97***<br>[0.867 - 1.074] | 3.08*<br>[-0.174 - 6.328] |
| Observations | 740 | 740 |
| R-squared | 0.014 | 0.101 |
| F-statistic | 3.546 | 4.725 |

Robust ci in brackets
\*\*\* p<0.01, \*\* p<0.05, \* p<0.1

Table A3 in the Appendix shows the results of the difference-in-differences analysis when considering the calendar period from 1 January to 31 August in each calendar year, thus including months after the end of the lockdown. This model confirms the findings of the baseline analysis, i.e. there is no relative increase in the number of suicides during the lockdown compared to what we would have expected in the absence of the pandemic.

## 4. Discussion

We studied suicide trends in Greece during the first wave of the Covid-19 pandemic and found no evidence of any increase after the introduction of the lockdown, which coincided with the period following the first covid-19 death, or during the period following the first wave, up to the end of August 2020. Our findings add to previous studies that did not find any increase in suicides in Japan **during the first wave** of the pandemic.^27,33^ We note, however, that the most recent reports indicate an increase in suicide, particularly among women, during the second wave of the pandemic in Japan.^26^

As discussed earlier, the effects of the pandemic on mental health and suicide rates would theoretically depend on the individual’s circumstances. Those who experienced job loss or lower incomes – or alternatively, daily worries due to being forced to continue to work in frontline occupations – would be expected to experience a deterioration in their mental health. Those who were able to switch to tele-work might have enjoyed a health dividend from avoiding commuting, and having more discretionary time (so long as they did not have young children to look after at home).^34^ The effects of lockdowns and physical distancing policies would themselves produce an increase in social isolation and loneliness. The **net effects** of the first wave of the Covid-19 pandemic on population suicide rates therefore remain a matter for empirical testing. In the case of Greece, at least during the initial wave of the pandemic, we did not observe any excess in suicide at the national level.

It is important to note that our results do not imply that suicides did not increase for vulnerable groups – e.g., those who lost their jobs or homes (due to falling behind on their rent), those who lost their loved ones, those who were lonely and isolated. or those who were forced to work in frontline occupations. Nor do our results imply that *mental health* did not deteriorate for the population as a whole. Suicide is just one particular outcome of poor mental health, and other aspects might have worsened (e.g. alcoholism), even if this did not translate into more suicides. In addition, our findings may not extend beyond the first wave of the pandemic, as the uncertainty becomes prolonged and economic hardship accumulates.

That said, our case study of Greece may hold lessons for factors that have played a protective role with regards to suicide rates in the country. First, Greece managed to keep the burden of Covid-19 cases to a relatively low level in the first wave,^30^ possibly as a result of a swift implementation of preventive measures.^35^ Secondly, Greece demonstrated a lower than-EU average rate of job loss according to Eurostat, with less than 2 percent of workers losing their job in the second quarter of 2020, as opposed to more than 6.5 percent in Spain; between 2 and 3.5 percent in France, Belgium, Netherlands, Sweden and Denmark; and between 3.5 and 5 percent in Italy, Portugal, Austria and Finland.^1^ However, over a quarter of workers in Greece faced reduced hours, which left Greece amongst the EU countries that scored the worst in this domain.^1^ Furthermore, universal health insurance applies in European countries, as opposed to the US, where workers who lose their job are also likely to lose their health insurance. Greece also demonstrated relatively strong family cohesion;^36^ advertising of suicide prevention hotlines; and income support policies.^35,37^ In addition to the 4-digit suicide prevention hotline by NGO Klimaka, the Ministry of Health also launched its own hotline during the lockdown, which was widely advertised. Things might deteriorate in the second wave, which saw a steep increase in confirmed Covid-19 cases, hospitalisations and deaths. This is something that future research can examine.

This study is subject to limitations. The dataset used included preliminary data available from the Suicide Prevention Observatory, rather than final official data. Furthermore, as the unit of observation was the total number of suicides per day, we could not account for individual characteristics, or for particular individual circumstances. In addition, we could not know whether reporting was in any way affected by the pandemic.

Most of the world is now going through the second wave of the pandemic. As the death toll increases, new lockdowns are being introduced and economic conditions worsen. Fatigue may be accumulating, while more people are also experiencing job loss or financial trouble, possibly resulting in even worse mental health outcomes that in the first wave. Particularly in Greece, the second wave is demonstrating a steep increase in confirmed cases and deaths,^[30]^ which could potentially lead to a totally different effect on suicides compared to the first wave. It is important that policy makers introduce relevant policies to respond appropriately to this mental health crisis facing our societies.^38^ These may include policies focusing directly on mental health,^39^ but also those targeting the root of the problem, such as unemployment and social safety nets.^40-41^ A 4-digit suicide prevention hotline in Greece might have played a protective role with regards to suicides during the first wave. Memorable numbers might be of particular help in times of crisis, and other countries could consider swiftly implementing such numbers (rather than longer numbers) in these times of crisis when mental health faces particular challenges.^42^

## Data Availability

Data availability statement: The data used on unemployment rates are freely available on the website of the Greek Statistics Authority. Suicide data were provided by the Suicide Prevention Centre, Klimaka NGO, Athens, Greece.

## Ethics approval

The data used were aggregate anonymous data so ethics approval was not required.

## Checklist

There is no relevant checklist of observational studies.

## Data availability statement

The data used on unemployment rates are freely available on the website of the Greek Statistics Authority. Suicide data were provided by the Suicide Prevention Centre, Klimaka NGO, Athens, Greece.

## APPENDIX

**Figure A1.**
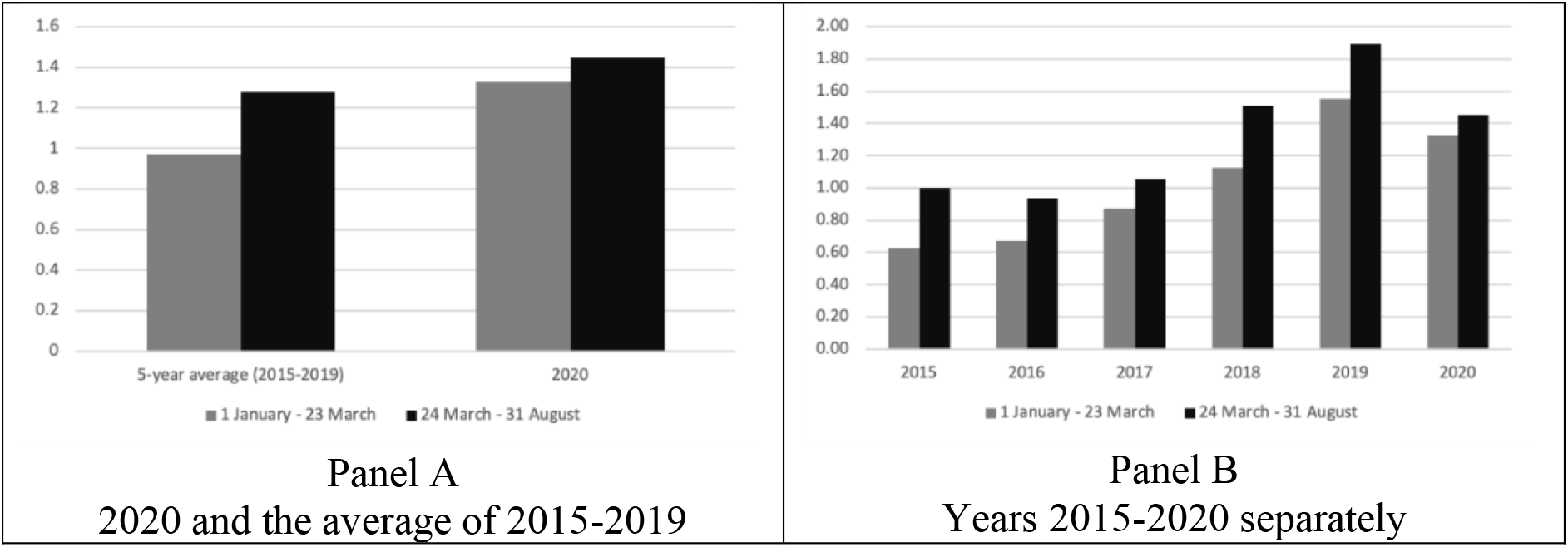
Average number of daily suicides before and after the introduction of the lockdown on 24 March 2020, until 31 August 2020 Data source: Suicide Observatory, Suicide Prevention Centre, Klimaka NGO

**Table A1.** Summary Statistics

| Variable | Mean | Std. Dev. | Min | Max |
| --- | --- | --- | --- | --- |
| Daily umber of suicides | 1.15 | 1.19 | 0 | 7 |
| Unemployment rate | 20.71 | 3.28 | 14.9 | 27.4 |
| Diff-in-diff coefficient | 0.08 | 0.27 | 0 | 1 |
| Treatment period | 0.76 | 0.42 | 0 | 1 |
| Treatment group | 0.12 | 0.32 | 0 | 1 |

**Table A2.**
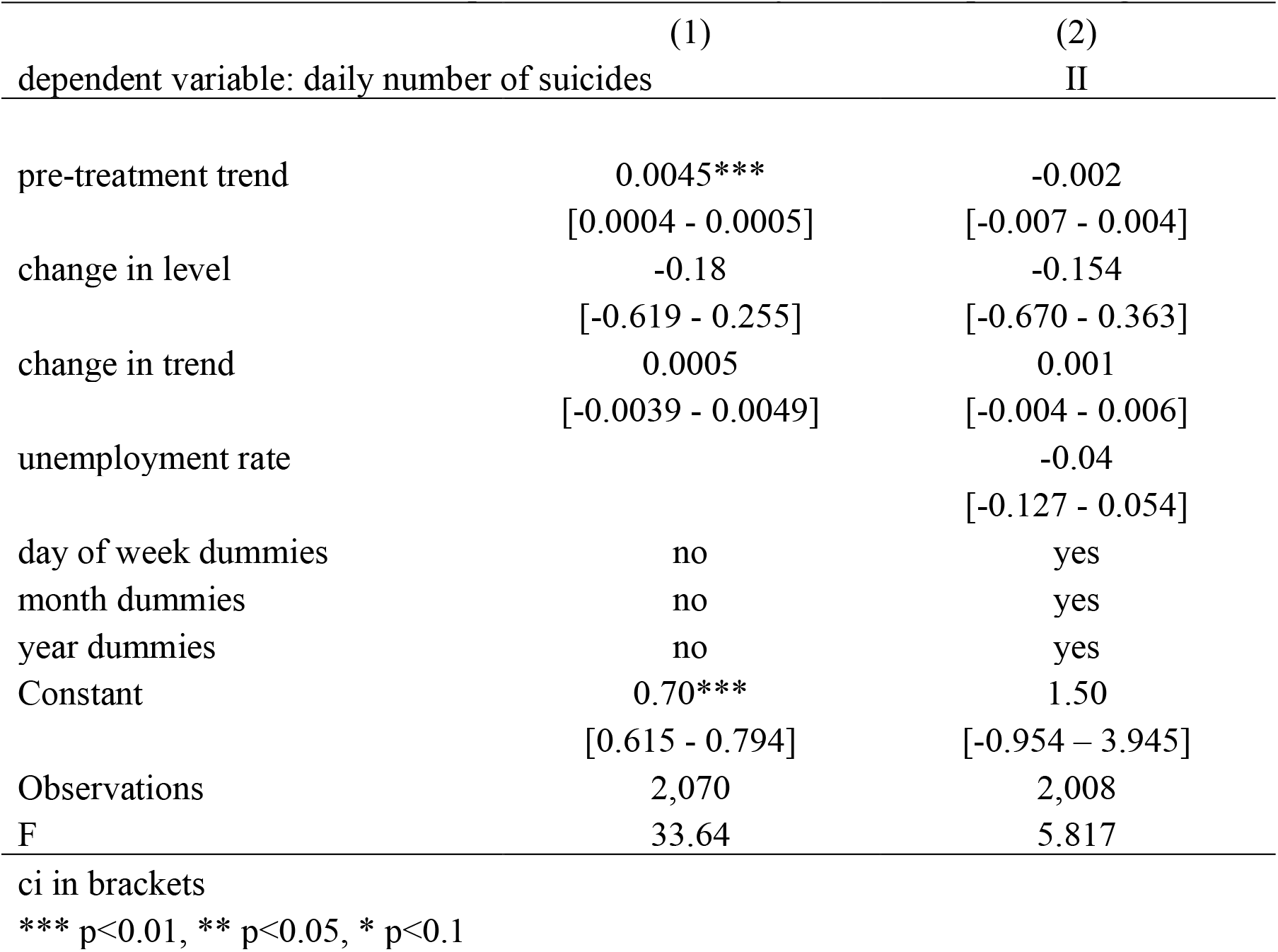
Results of the Interrupted Time Series analysis. Period up to 31 August 2020

|  | (1) | (2) |
| --- | --- | --- |
| dependent variable: daily number of suicides |  | II |
| pre-treatment trend | 0.0045***<br>[0.0004 - 0.0005] | -0.002<br>[-0.007 - 0.004] |
| change in level | -0.18<br>[-0.619 - 0.255] | -0.154<br>[-0.670 - 0.363] |
| change in trend | 0.0005<br>[-0.0039 - 0.0049] | 0.001<br>[-0.004 - 0.006] |
| unemployment rate |  | -0.04<br>[-0.127 - 0.054] |
| day of week dummies | no | yes |
| month dummies | no | yes |
| year dummies | no | yes |
| Constant | 0.70***<br>[0.615 - 0.794] | 1.50<br>[-0.954 - 3.945] |
| Observations | 2,070 | 2,008 |
| F | 33.64 | 5.817 |

ci in brackets
\*\*\* p<0.01, \*\* p<0.05, \* p<0.1

**Table A3.**
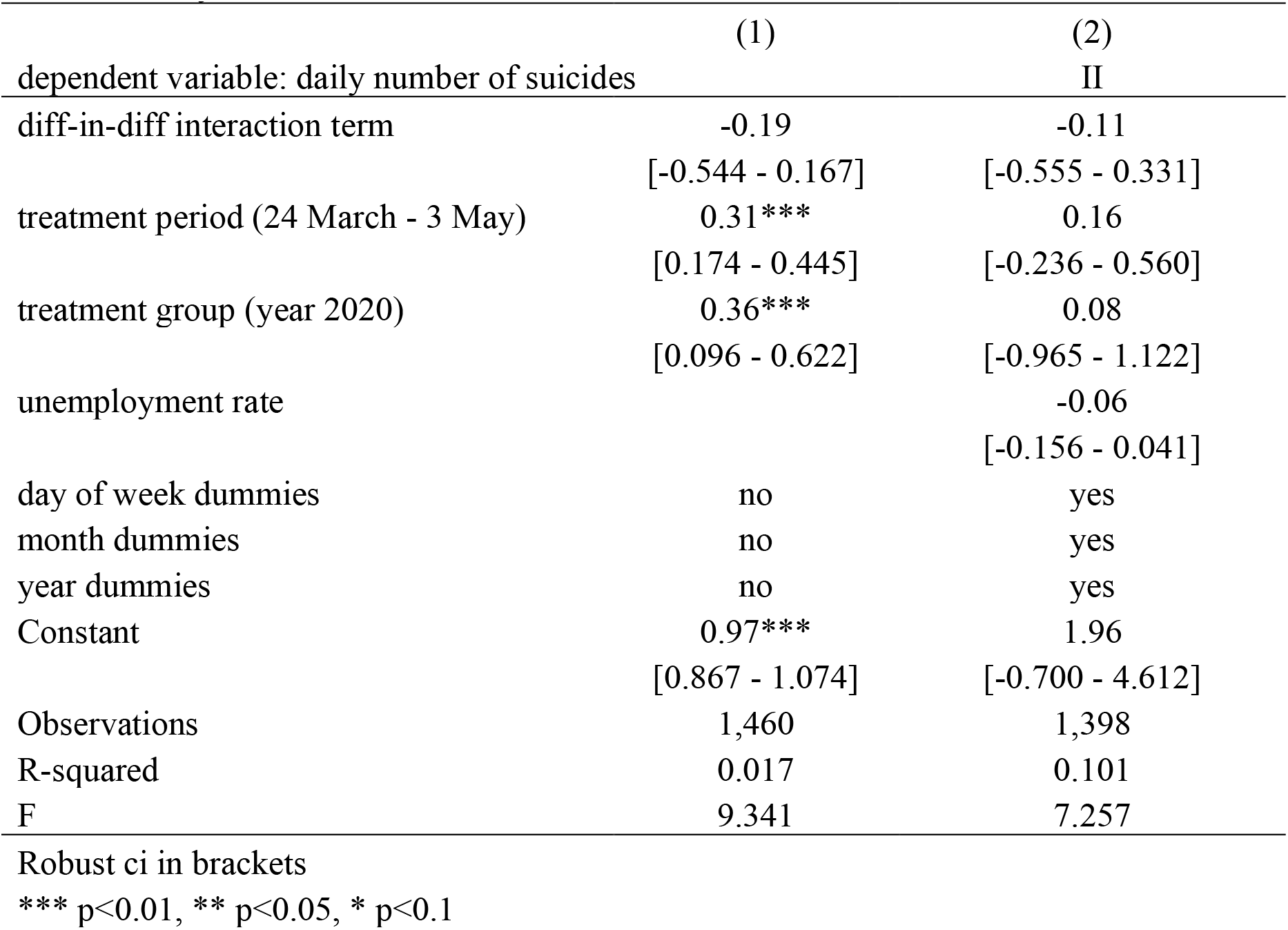
Results of the differences-in-differences model. Period 1 January – 31 August of each calendar year

|  | (1) | (2) |
| --- | --- | --- |
| dependent variable: daily number of suicides |  | II |
| diff-in-diff interaction term | -0.19<br>[-0.544 - 0.167] | -0.11<br>[-0.555 - 0.331] |
| treatment period (24 March - 3 May) | 0.31***<br>[0.174 - 0.445] | 0.16<br>[-0.236 - 0.560] |
| treatment group (year 2020) | 0.36***<br>[0.096 - 0.622] | 0.08<br>[-0.965 - 1.122] |
| unemployment rate |  | -0.06<br>[-0.156 - 0.041] |
| day of week dummies | no | yes |
| month dummies | no | yes |
| year dummies | no | yes |
| Constant | 0.97***<br>[0.867 - 1.074] | 1.96<br>[-0.700 - 4.612] |
| Observations | 1,460 | 1,398 |
| R-squared | 0.017 | 0.101 |
| F | 9.341 | 7.257 |

Robust ci in brackets
\*\*\* p<0.01, \*\* p<0.05, \* p<0.1

